# Healthcare workers’ COVID-19 Omicron variant uncertainty-related stress, resilience, and coping strategies during the first week of World Health Organization alert

**DOI:** 10.1101/2021.12.24.21268377

**Authors:** Mohamad-Hani Temsah, Shuliweeh Alenezi, Mohamad Al-Arabi, Fadi Aljamaan, Khalid Alhasan, Rasha Assiri, Rolan Bassrawi, Fatimah Alshahrani, Ali Alhaboob, Ali Alaraj, Naser S Alharbi, Rabih Halwani, Amr Jamal, Naif Abdulmajeed, Lina Alfarra, Wafa Almashdali, Ayman Al-Eyadhy, Fahad AlZamil, Sarah Al-Subaie, Mazin Barry, Ziad A. Memish, Jaffar A. Al-Tawfiq

## Abstract

**Background:** As the COVID-19 Omicron variant emerged and spread globally at an alarming speed, healthcare workers’ (HCWs) uncertainties, worries, resilience, and coping strategies warrant assessment. The COVID-19 pandemic had a severe psychological impact on HCWs, including the development of Post-Traumatic Stress symptoms. Specific subgroups of HCWs, such as front-line and female workers, were more prone to poor mental health outcomes and difficulties facing stress.

**Methods:** The responses to an online questionnaire among HCWs in Saudi Arabia (KSA) were collected December 1-5, 2021, aiming to assess their Omicron variant’s uncertainties, worries, resilience, and coping strategies. Three validated instruments were used to achieve the study’s goals: the Brief Resilient Coping Scale, the Standard Stress Scale (SSS), and the Intolerance of Uncertainty Scale (IUS) - Short Form.

**Results:** The online survey was completed by 1285 HCWs. Females made up the majority (64%). The BRCS score of resilient coping was negatively and substantially linked with the SSS score of stress (r=-0.313, p = 0.010). Furthermore, the IUS had a positive and significant relationship with stress (r=0.326, p= 0.010). Increased stress levels were linked to a considerable drop in resilient coping scores. Furthermore, being a Saudi HCW or a nurse was linked to a significant reduction in resilient coping ratings. Coping by following healthcare authorities’ preventative instructions and using the WHO website as a source of information was linked to a considerable rise in resilient coping.

**Conclusions:** Following the emergence of the Omicron variant of SARS-CoV-2 in late 2021, a rapid investigation into the correlates of stress and resilient coping among the HCWs in KSA was conducted. The negative association between resilient coping and stress was clearly shown, as well as how underlying intolerance of uncertainty is linked to higher stress among HCWs quickly following the development of a new infectious threat. The study provides early insights to develop and promote coping strategies for emerging SARS-CoV-2 variants.

## 1 Introduction

In November 2021, researchers in South Africa announced the emergence of a new variant of SARS-CoV-2[1]. Later, the World Health Organization (WHO) designated this variant as a variant of concern and named it Omicron.[2] The appearance of a new infectious threat presented healthcare workers (HCWs) with a new source of stress and worry.

Previous research has shown that the COVID-19 pandemic had a significant psychological toll on HCWs [3-9], including the development of Post-Traumatic Stress symptoms[10,11]. Certain subgroups within HCWs, such as front-line and female workers, were particularly vulnerable to worse mental health outcomes[12-15]. Furthermore, a study on healthcare workers following the spread of the SARS-CoV-2 Delta variant revealed high levels of worry[16].

HCWs have been struggling with uncertainty since the beginning of the COVID-19 pandemic [17]. This uncertainty was not limited to the possibility of infection but extended to the possible socio-economic impact of the pandemic[18]. Notably, difficulty in tolerating uncertainty may underlie the worry they experience in stressful situations [19]. Indeed, the construct of Intolerance of Uncertainty was developed to capture this tendency [20], which also contributes to the development of anxiety disorders [21]. In the context of the COVID-19 pandemic, higher intolerance of uncertainty has been found to correlate with stress and anxiety in various countries during the pandemic [22-30]. A similar correlation was also observed in HCWs, whose intolerance of uncertainty was correlated with their utilized coping strategies in the face of the pandemic[31]. Thus, it is crucial to study HCWs’ intolerance of uncertainty, and a vital construct for additional studies as the uncertainties of the pandemic persists. This is particularly important with the emergence of the Omicron variant, with its many unknowns appearing in the public health scene [1].

An essential contributor to the mental wellbeing of HCWs is their ability to cope with the continuing stress of the pandemic [32,33] and their perceived resilience [34,35]. This is most pertinent with the emergence of new variants that can threaten their health and lead to more stressful working environments. Most relevantly, HCWs with lower resilience and higher intolerance of uncertainty were at higher risk of developing burnout during the COVID-19 pandemic [36]. Since HCWs’ stress and subsequent burnout may increase with time during the pandemic [37], the appearance of the Omicron variant may further increase stress and burnout by lengthening the pandemic for months or years.

On December 1^st^, 2021, The Saudi Ministry of Health (MoH) announced the detection of the first case of the omicron variant in the country [38]. With that news present in the consciousness of many HCWs, we aimed to assess their awareness and sources of worry in relation to the Omicron variant and how these correlate with their perceived stress and ability to cope with stressful situations. Our goals also included exploring how intolerance of uncertainty relates to stress and whether higher resilient coping was correlated with decreased stress.

## 2 Method

This was a national, cross-sectional survey among HCWs in KSA that was conducted between December 1 and 6, 2021. At that period, several countries had reported infection with the new SARS-CoV-2 Omicron variant, and KSA reported only one case. HCWs were invited through convenience sampling technique by several professional social media platforms, including WhatsApp groups and Twitter posts, and email lists. Participants were asked about their Omicron variant awareness, worry, and stress with the emergence of the quickly spreading Omicron variant and their resilience and coping strategies during the pandemic crisis. The survey was pilot-validated and electronically distributed through SurveyMonkey^©^. The questionnaire was adapted from our previously published studies on COVID-19 stress and coping, with modifications related to the new SARS-CoV-2 Omicron variant[16,39-42].

To assess HCWs’ perceived resilience, we incorporated the Brief Resilient Coping Scale (BRCS), a reliable and valid tool for self-rated assessment of resilient coping[43]. The scale items had good internal consistency in our sample (Cronbach’s Alpha = 0.85). In addition, to measure HCWs’ stress levels, we utilized a self-report scale that was developed to assess stress in various circumstances and for a range of demographics. The Standard Stress Scale (SSS) is a 5-point Likert scale composed of 11 items that measure stress[44]. The scale has good psychometric properties, and our sample had good internal consistency (Cronbach’s Alpha= 0.70). Finally, we incorporated the Intolerance of Uncertainty Scale (IUS) to quantify our respondents’ underlying intolerance of uncertainty and relate it to their stress levels[45]. The IUS-12 is a briefer form of the original scale of 12 5-point Likert items. The scale has good psychometric properties, including our sample of HCWs (Cronbach’s alpha = 0.89).

The questions about HCWs’ demographics included job category, age, sex, and work area, previous exposure to COVID-19 patients in the last three months, whether the HCW was previously infected with COVID-19 themselves, travel history to a country with the Omicron variant in the previous one month, and the COVID-19 vaccines they received. We assessed factors affecting HCWs’ worry level regarding international travel and their sources of information about the SARS-CoV-2 variants. HCWs’ anxiety was also measured by asking them to self-rate their worry levels on a 5-items Likert scale, comparing the worry towards the original COVID-19 strain, the Alpha, the Delta, and the Omicron variants.

### 2.1 Data Collection

Participants were informed before starting the survey of the purpose of this study and that their participation in this research was completely voluntary. The Institutional Review Board at King Saud University approved the study (approval 21/01039/IRB).

### 2.2 Statistical analysis

The mean and standard deviation were used to describe the continuous variables, and the frequency and percentage to describe the categorically measured variables. The histogram and the K-S statistical test of Normality were used to assess the statistical Normality assumption of the continuous variables, and Levene’s test was used to assess the homogeneity of variance statistical assumption. The reliability analysis of the measured psychometric scales was tested with the Cronbach’s alpha test. The multiple response dichotomies analysis was used to analyze the multiple response variables. The Pearson’s correlations test (r) was used to assess the correlations between metric variables. The Multivariable Linear Regression Analysis was applied to assess the statistical significance of the predictors of HCW’s perceived stress and resilient coping scores, and the tested predictor independent variables were selected based on their (theoretical, practicable, managerial or statistical) relevance. The association between these predictors with the analyzed dependent outcome variables was expressed as unstandardized beta (β) coefficients with their 95% confidence intervals. The SPSS IBM statistical analysis program Version#21 was used for the statistical data analysis. The statistical significance level was considered with P value of ≤ 0.05.

## 3 Results

A total of 1285 HCW’s completed the online survey, from all regions in KSA. Most of HCW’s (57.6% were working in Riyadh Capital City and the central region. Most of the participants (64%) were females and expatriates (62.3%). Their age distribution is shown in table 1. Most of HCW’s (49.8%) were nurses and (24.8%) were medical consultants. Almost half of responders (50.2%) worked in Tertiary health centers.

**Table.1:** Descriptive analysis of the HCW’s sociodemographic characteristics and professional attributes. N=1285.

|  | Frequency | Percentage |
| --- | --- | --- |
| **Gender** |  |  |
| Female | 822 | 64 |
| Male | 463 | 36 |
| **Age group (in years)** |  |  |
| 25-34 | 434 | 33.8 |
| 35-44 | 477 | 37.1 |
| 45-54 | 273 | 21.2 |
| ≥ 55 years | 101 | 7.9 |
| **Nationality** |  |  |
| Saudi | 484 | 37.7 |
| Expatriate | 801 | 62.3 |
| **Clinical Role** |  |  |
| Consultant | 319 | 24.8 |
| Assistant consultant / Fellow | 74 | 5.8 |
| Resident / Registrar | 203 | 15.8 |
| Nurse | 640 | 49.8 |
| Technician | 49 | 3.8 |
| **Hospital type** |  |  |
| Primary healthcare center | 338 | 26.3 |
| Secondary hospital | 302 | 23.5 |
| Tertiary hospital | 645 | 50.2 |
| **Hospital working area** |  |  |
| Intensive care units (ICU) | 141 | 11 |
| Emergency Room (ER) | 91 | 7.1 |
| Operating Room (OR) | 41 | 3.2 |
| COVID-19 Isolation ward | 53 | 4.1 |
| General ward | 492 | 38.3 |
| Outpatient Department (OPD) | 368 | 28.6 |
| Non-clinical area | 99 | 7.7 |
| **Region** |  |  |
| Central region | 740 | 57.6 |
| Eastern Provinces | 71 | 5.5 |
| Western Provinces | 120 | 9.3 |
| Northern Provinces | 34 | 2.6 |
| Southern Provinces | 320 | 24.9 |

Of the respondents, 38.3% worked in General Hospital Wards, and 11%, 7.1% and 4.1% worked in intensive care units (ICU), emergency room (ER) and COVID-19 Isolation wards, respectively.

Table 2 displays the findings of the surveyed HCW’s experiences about COVID-19 disease. Of the HCWs, 29% reported recent contact with COVID-19 patients, while 22.3% were previously diagnosed as COVID-19 themselves. Over the past two years, the mean number of PCR testing performed per HCWs due to suspicion of SARSCoV-2 infection was (3.43 + 3.2 times). Only 2.1% of the HCW’s had been to countries with identified Omicron variant spread in the last month.

**Table.2:** Descriptive analysis of the HCW’s experiences of COVID-19 disease, screening, and immunization.

|  | Frequency | Percentage |
| --- | --- | --- |
| **In the past 3 months: Have you been in contact with COVID-19 patients?** |  |  |
| No | 912 | 71 |
| Yes | 373 | 29 |
| **Were you previously diagnosed with PCR-positive COVID-19 yourself?** |  |  |
| No | 999 | 77.7 |
| Yes | 286 | 22.3 |
| **How many COVID-19 tests have you had since the pandemic started?**<br>**Mean (SD)** |  | 3.43 (3.20) |
| **During the last month: Did you travel to any country where the Omicron variant has been recorded?** |  |  |
| Yes | 27 | 2.1 |
| No | 1258 | 97.9 |
| **How familiar are you with the new Omicron variant? Mean (SD)** |  | 3.24 (0.95) |
| Extremely aware | 132 | 10.3 |
| Very aware | 329 | 25.6 |
| Somewhat aware | 579 | 45.1 |
| Not so aware | 199 | 15.5 |
| Not at all aware | 46 | 3.6 |
| **How familiar are you with the Delta variant? Mean (SD)** |  | 3.50 (0.99) |
| Extremely aware | 204 | 15.9 |
| Very aware | 452 | 35.2 |
| Somewhat aware | 443 | 34.5 |
| Not so aware | 149 | 11.6 |
| Not at all aware | 37 | 2.9 |
| **Using a Likert rating from 1-5, How worried are you from** |  |  |
| From International travel. Mean (SD) |  | 3.19 (1.12) |
| The original strain that started the first pandemic. Mean (SD) |  | 1.96 (1.14) |
| The Alpha variant (that was first described in the UK). Mean (SD) |  | 1.67 (1.1) |
| The Delta variant (that was first described in India). Mean (SD) |  | 1.97 (1.13) |
| The new Omicron variant. Mean (SD) |  | 2.18 (1.14) |

HCW’s were asked to self-rate their familiarity level with the Omicron and the Delta variants, the mean familiarity score was 3.24 out of 5-Likert’s points with Omicron variant and 3.5 for the Delta variant. As shown in figure 2, the top accessed source of information by the HCW’s was the WHO website (51.5%), followed by the Saudi MoH website (50.4%) and the Social Media channels and news (41%). In addition, 38%followed information released by formal spokespeople, and 33.5% relied on hospital announcements and the Saudi Center for Disease Control CDC website information as well as the US CDC website. Of the responders, 28.7% had learnt about the Omicron variant from medical Journals, and 16% from other sources/channels of information. HCW’s were asked to self-rate their worry level from international travel and the various SARS-CoV-2 variants. The mean worry from travel abroad was 3.19/5 points, the worry from the original SARS-COV-2 variant was 1.96/5 points, and from the Alpha variant was 1.67/5 points; however, the Delta variant worry level was 1.97/5and from the Omicron variant was 2.18/5 points.

**Figure 1:**
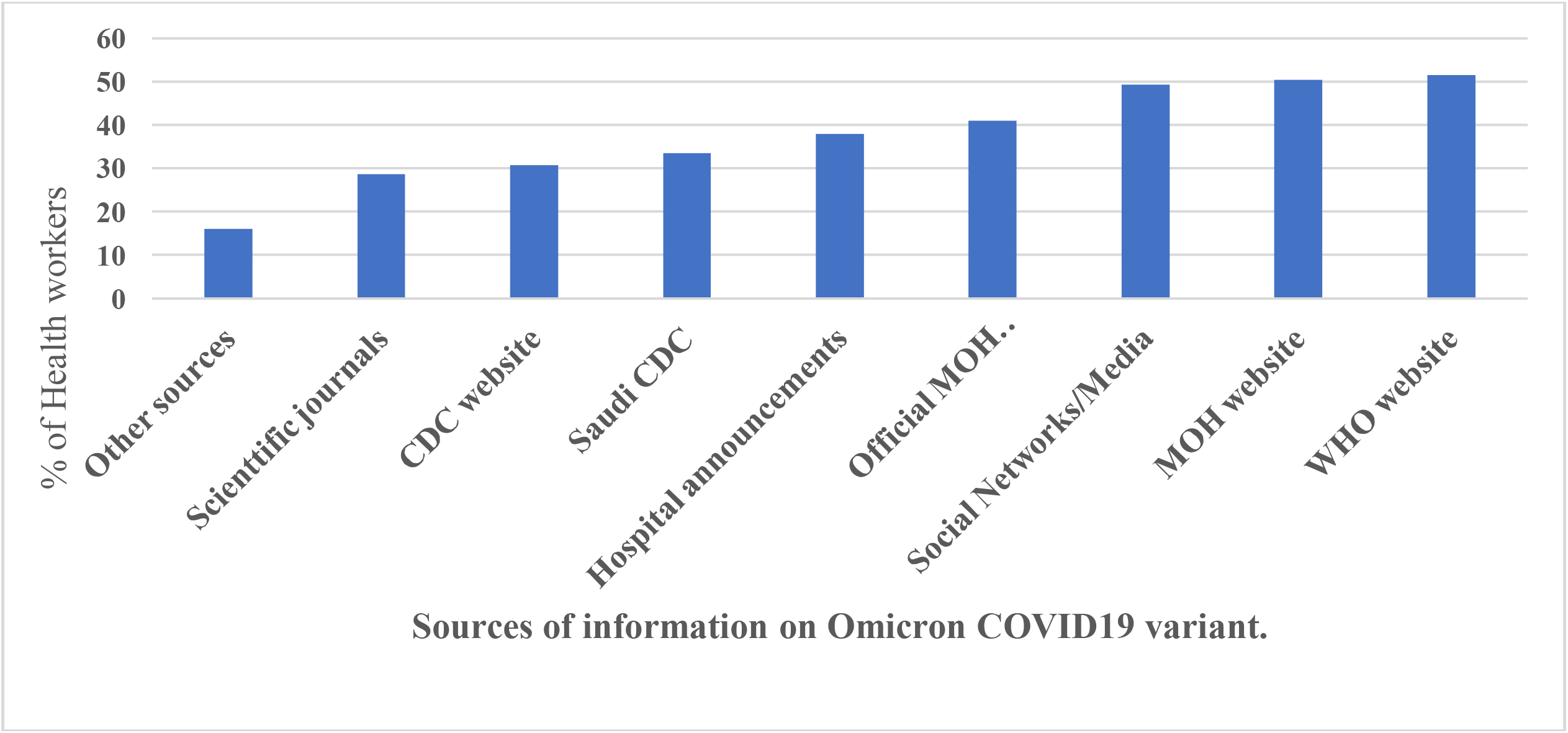
The HCWs’ used sources of information on the Omicron variant.

**Figure 2:**
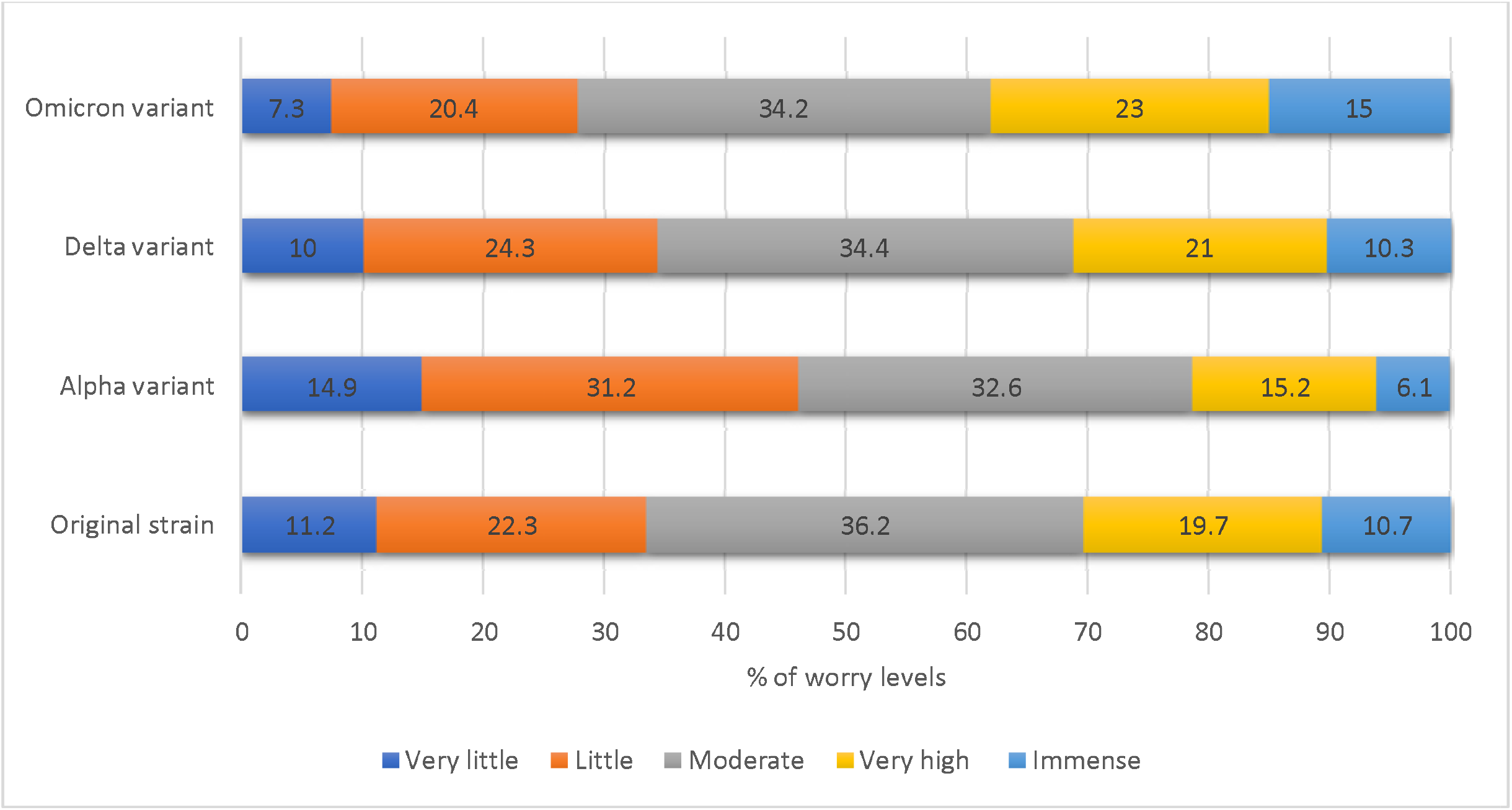
The HCWs’ perceived worry levels from various SARS-CoV-2 strains *. * Worries from original, alpha, and delta strains were significantly lower than the worries from Omicron (paired samples t-test, p<0.001 each respectively)

Table 3A shows the mean, standard deviation, and rank of the means for the surveyed HCW’s perceptions of their resilient coping, stress, and intolerance of uncertainty. The top perceived coping indicator according to the BRCS in our sample was the ability to grow in positive way by handling difficulties, followed by active looking for ways to replace the losses encountered in life, then the ability to control reactions to various situations and lastly, looking for creative ways to adjust challenging situations.

**Table.3:** Descriptive analysis of the HCWs’ stress, resilient coping and intolerance of uncertainty.

|  | **Mean** | **SD** | **Possible score range** | **Equivalent Percentage for the Mean** |
| --- | --- | --- | --- | --- |
| Standard Stress Scale (SSS) score | 0.42 | 0.11 | 0-1 points | 42.0 |
| Intolerance of Uncertainty (IUS-12) score | 31.81 | 8.52 | 12-60 points | 53.0 |
| Brief Resilient Coping Scale (BRCS) score | 14.31 | 2.88 | 4-20 points | 71.6 |
| Resilient coping level | Frequency | Percentage |  |  |
| Low resilient coping | 509 | 39.6 | — |  |
| Medium resilient coping | 541 | 42.1 | — |  |
| High resilient coping | 235 | 18.3 | — |  |

The surveyed HCWs ranked their worries from original, alpha, and delta strains as significantly lower than the worries from the emerging Omicron (paired samples t-test, p<0.001, Figure 2). Regarding the perceived stress indicators as assessed by the SSS, doing meaningful tasks was on the top of the list, then looking forward to the future, and having people around that they could count on. Conversely, the lowest respondents perceived stress indicators were being afraid from how their life will look like in three years, feeling exhausted after normal working days and having restorative sleep.

The surveyed HCW’s top perceived indicator of uncertainty as scored by the IUS-12 was their ability to organize everything in their life ahead of time, this was followed by their agreement with that one should always look ahead to avoid surprise. The 3rd top indicator was feeling frustrated if they do not have all the information they need and lastly, was their constant willingness to know what the future hides for them. However, the lowest rated perceptions of uncertainty for respondents were being stopped from action by smallest doubts then being paralyzed by uncertainty when action is needed in their life (table 2).

The overall mean perceived SSS score was 0.42 /1 points, SD= 0.11 points and IUS-12 score was 31.81/60 points, SD=8.52 points. Whereas the mean BRCS total score was 14.31/20 points, SD= 2.88 points. If cut-off scores were used for the BRCS, 39.6% of HCWs were considered to have low resilient coping and only 18% were considered to have high resilient coping (table 3).

For most of surveyed HCW’s, the risk of transmitting the infection to home and new lockdown or disruption of normal daily life were the top perceived source of concern about the Omicron viral outbreak followed by new travel ban concern (table 4). The higher risk of Omicron transmissibility was a concern for 57.1%. Overwhelmed healthcare services and lack of some equipment during the pandemic account for (46.3%) and (32.5%) of HCW’s source of worries.

**Table.4:** Descriptive analysis of the HCW’s beliefs, attitudes and practices concerning Omicron variant.

| Variables | Frequency | Percentage |
| --- | --- | --- |
| **Sources of worries and fears about SARS-CoV-2 Omicron variant** |  |  |
| Risk of taking the disease to my family and household | 791 | 63.2 |
| Risk of lockdown or disruption to normal daily life | 790 | 63.1 |
| Risk of travel ban | 765 | 61.1 |
| The higher risk of possible transmission among HCWs | 715 | 57.1 |
| Overwhelmed healthcare services | 580 | 46.3 |
| Lack of some equipment during pandemic (example: ventilator shortages) | 407 | 32.5 |
| The risk of depletion of my hospital's PPE | 387 | 30.9 |
| Other worries | 97 | 7.7 |
| **Your best used coping strategies with Omicron as a HCW** |  |  |
| Following the prevention guidelines of healthcare authorities | 824 | 65.8 |
| Social distancing | 709 | 56.6 |
| Family support | 708 | 56.5 |
| Having faith | 684 | 54.6 |
| Seeking reliable information | 654 | 52.2 |
| Focusing on work | 523 | 41.8 |
| Sport and exercise | 481 | 38.4 |
| Seeking support from friends and colleagues | 415 | 33.1 |
| Reading | 409 | 32.7 |
| Staying at home | 400 | 31.9 |
| Avoiding reading about or discussing new SARS-CoV-2 strains | 157 | 12.5 |
| Other coping methods | 74 | 5.9 |

The respondents’ top used coping method was following the disease transmission prevention guidelines (65.8%) followed by applying social distancing (56.6.8%). Family support and bonding was helpful for (56.5%) as well as having faith (54.6%) and focusing on work (41.8%). For (52.2%) HCW’s, seeking reliable information about the disease is helpful for coping. However, few (12.5%) prefer to Avoid reading about or discussing new COVID-19 strains.

Table 5 shows the bivariate Pearson’s correlation coefficients between measures of resilient coping, stress, intolerance of uncertainty of HCWs, and their self-reported levels of worry and familiarity with current and past variants of COVID-19. Resilient coping as scored by the BRCS was negatively and significantly correlated with stress as scored by the SSS (r=-0.313, p<0.010). Moreover, Intolerance of Uncertainty as scored by the IUS correlated positively and significantly with stress (r=0.326, p<0.010). The analysis also revealed that self-rated worry levels were mostly correlated significantly but weakly with IUS and SSS scores. However, self-reported levels of worry regarding different aspects of current or past variants of COVID-19 correlated substantially with each other (r>0.50. p-value <0.010).

**Table.5:**
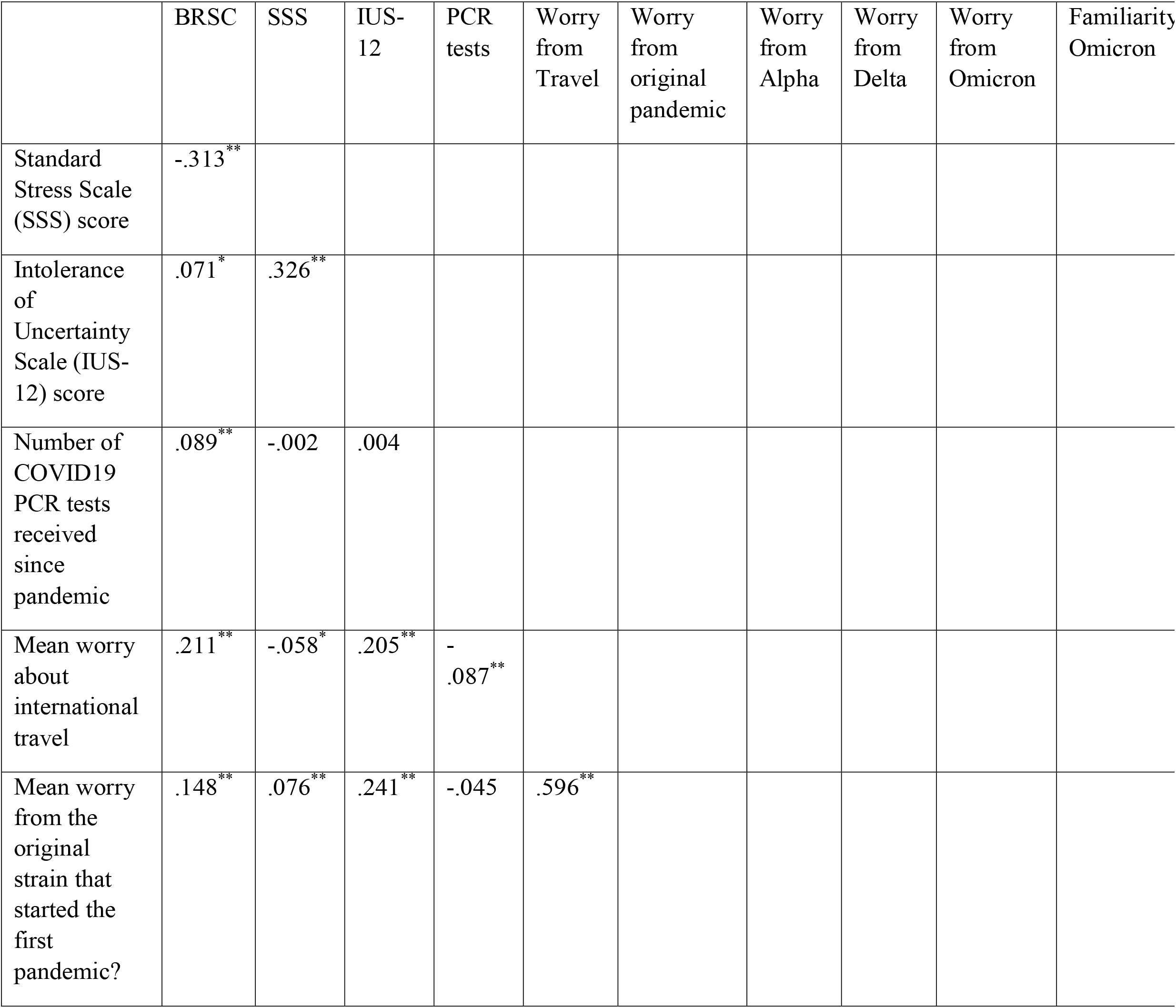
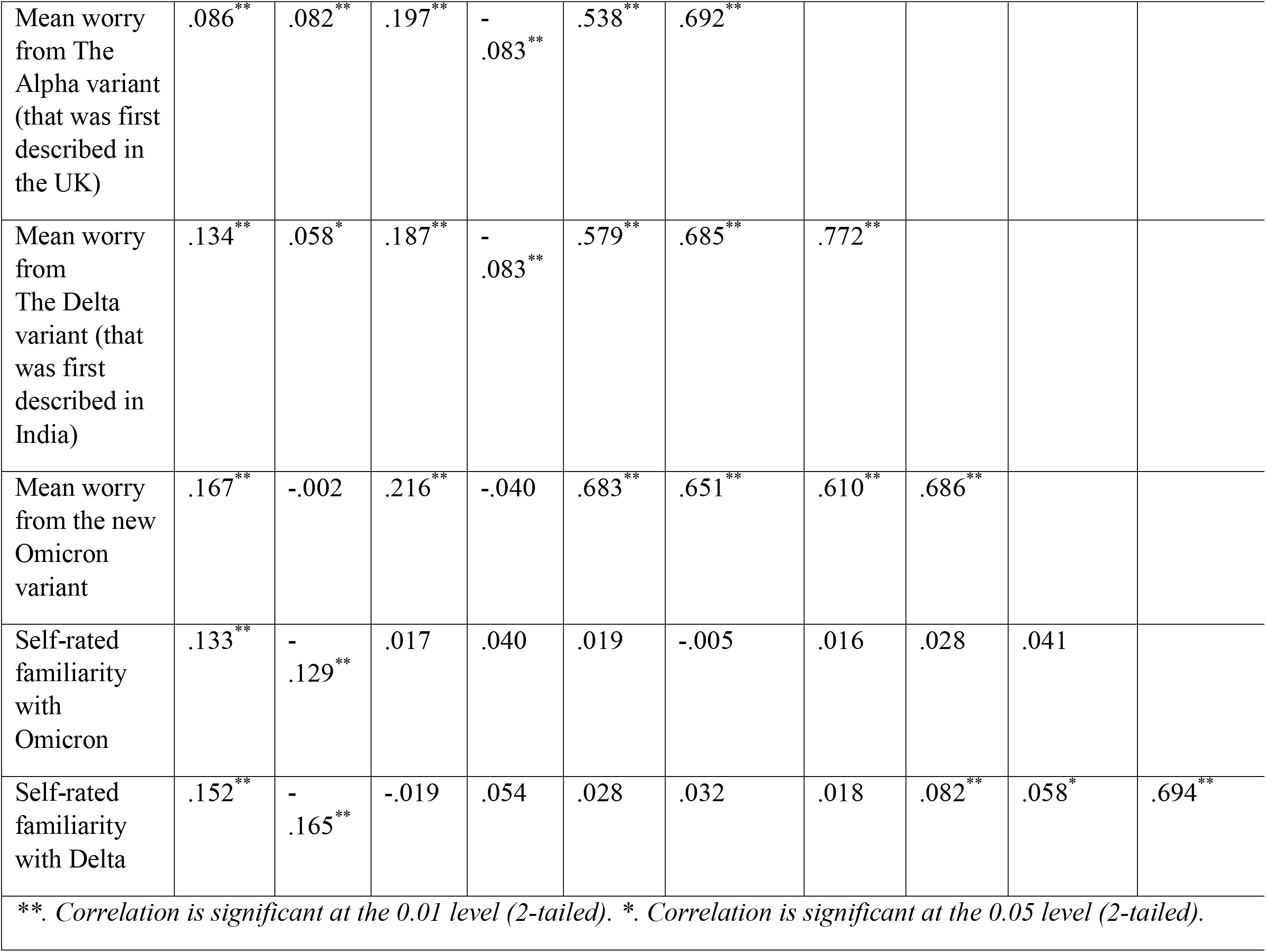
Bivariate Correlations between the HCW’s measured perceptions.

In order to explain the variability in reported resilient coping, we built a multiple linear regression model for BRCS scores using relevant variables (Table 6). The model is detailed in Table 7 and explained a significant amount of variation in resilient coping scores between HCWs (*adjusted R-squared=0*.*21, p<0*.*001)*. According to the model, the most significant association with resilient coping scores was stress scores, the higher the stress score the lower the coping score. In addition, being a Saudi or a nurse, both correlated with a significant decrease in resilient coping scores. On the other hand, coping by following the prevention guidelines of healthcare authorities and using the WHO website as a source of information were both associated with a significant increase in resilient coping.

**Table.6:**
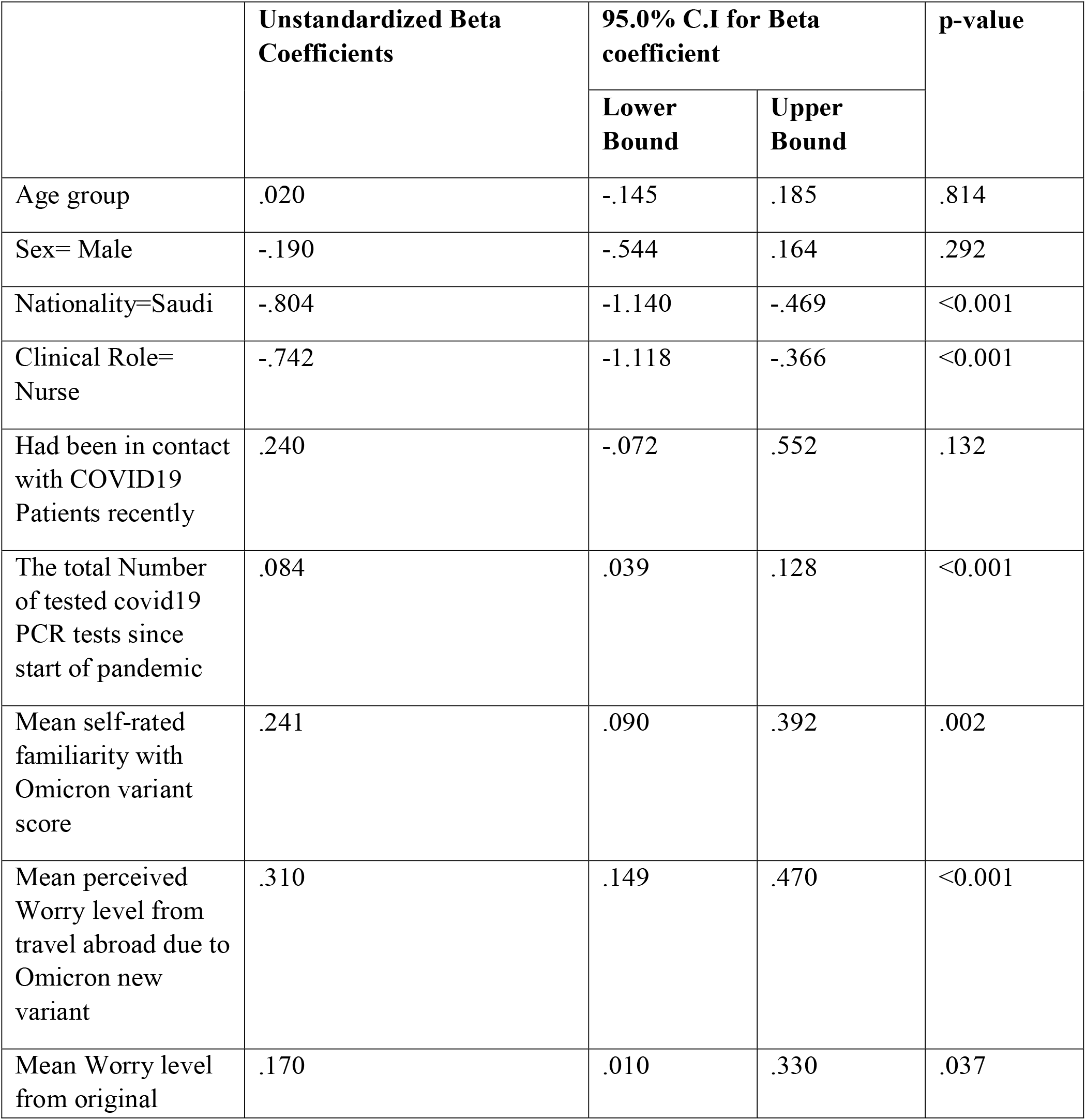
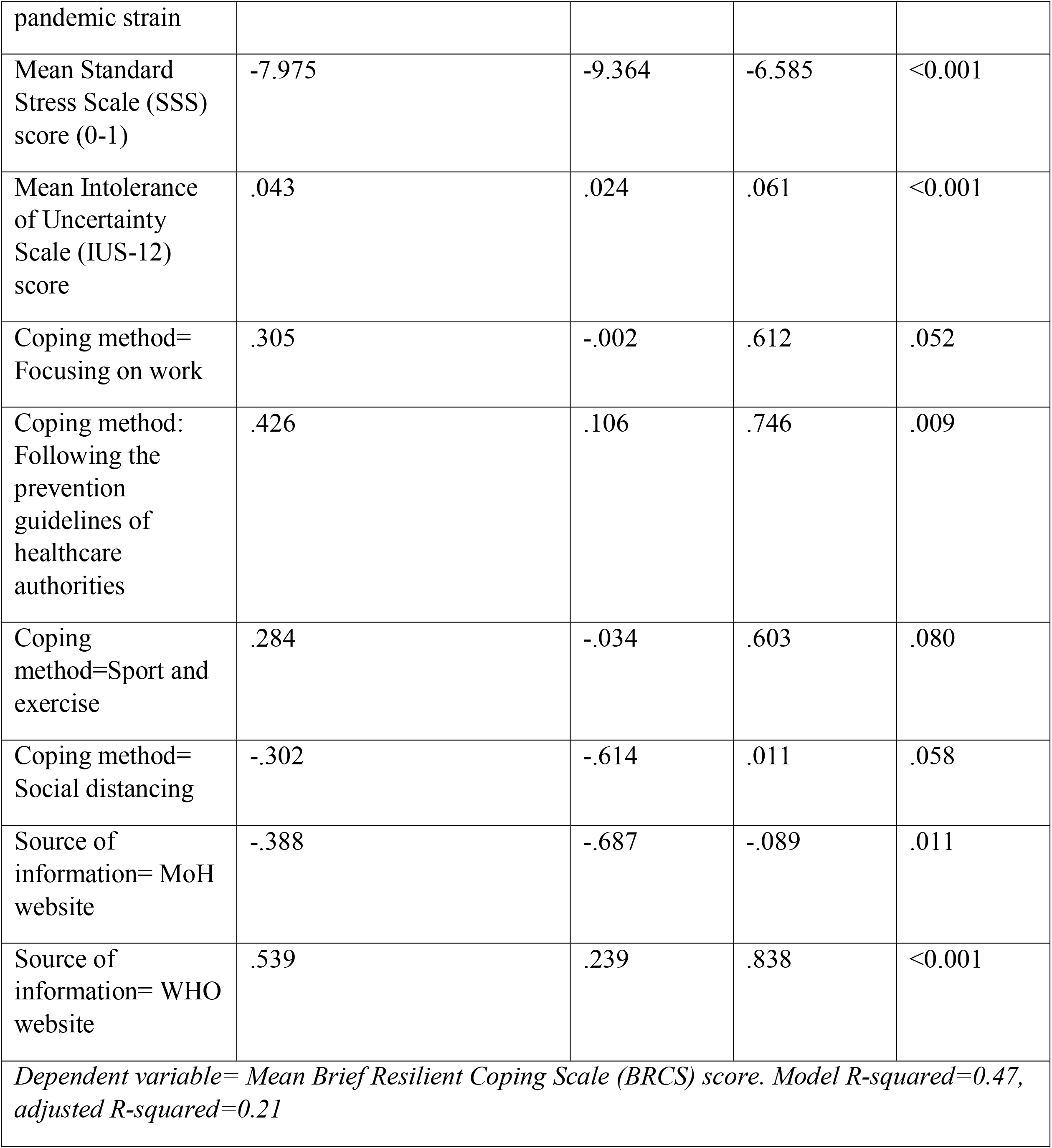
Multivariate Linear Regression Analysis of the HCWs’ resilient coping. N=1285.

|  | Unstandardized Beta Coefficients | 95.0% C.I for Beta coefficient |  | p-value |
| --- | --- | --- | --- | --- |
|  |  | Lower Bound | Upper Bound |  |
| Age group | .020 | -.145 | .185 | .814 |
| Sex= Male | -.190 | -.544 | .164 | .292 |
| Nationality=Saudi | -.804 | -1.140 | -.469 | <0.001 |
| Clinical Role= Nurse | -.742 | -1.118 | -.366 | <0.001 |
| Had been in contact with COVID19 Patients recently | .240 | -.072 | .552 | .132 |
| The total Number of tested covid19 PCR tests since start of pandemic | .084 | .039 | .128 | <0.001 |
| Mean self-rated familiarity with Omicron variant score | .241 | .090 | .392 | .002 |
| Mean perceived Worry level from travel abroad due to Omicron new variant | .310 | .149 | .470 | <0.001 |
| Mean Worry level from original | .170 | .010 | .330 | .037 |
| pandemic strain |  |  |  |  |
| Mean Standard Stress Scale (SSS) score (0-1) | -7.975 | -9.364 | -6.585 | <0.001 |
| Mean Intolerance of Uncertainty Scale (IUS-12) score | .043 | .024 | .061 | <0.001 |
| Coping method= Focusing on work | .305 | -.002 | .612 | .052 |
| Coping method: Following the prevention guidelines of healthcare authorities | .426 | .106 | .746 | .009 |
| Coping method= Sport and exercise | .284 | -.034 | .603 | .080 |
| Coping method= Social distancing | -.302 | -.614 | .011 | .058 |
| Source of information= MoH website | -.388 | -.687 | -.089 | .011 |
| Source of information= WHO website | .539 | .239 | .838 | <0.001 |
| *Dependent variable= Mean Brief Resilient Coping Scale (BRCS) score. Model R-squared=0.47, adjusted R-squared=0.21* |  |  |  |  |

**Table.7:**
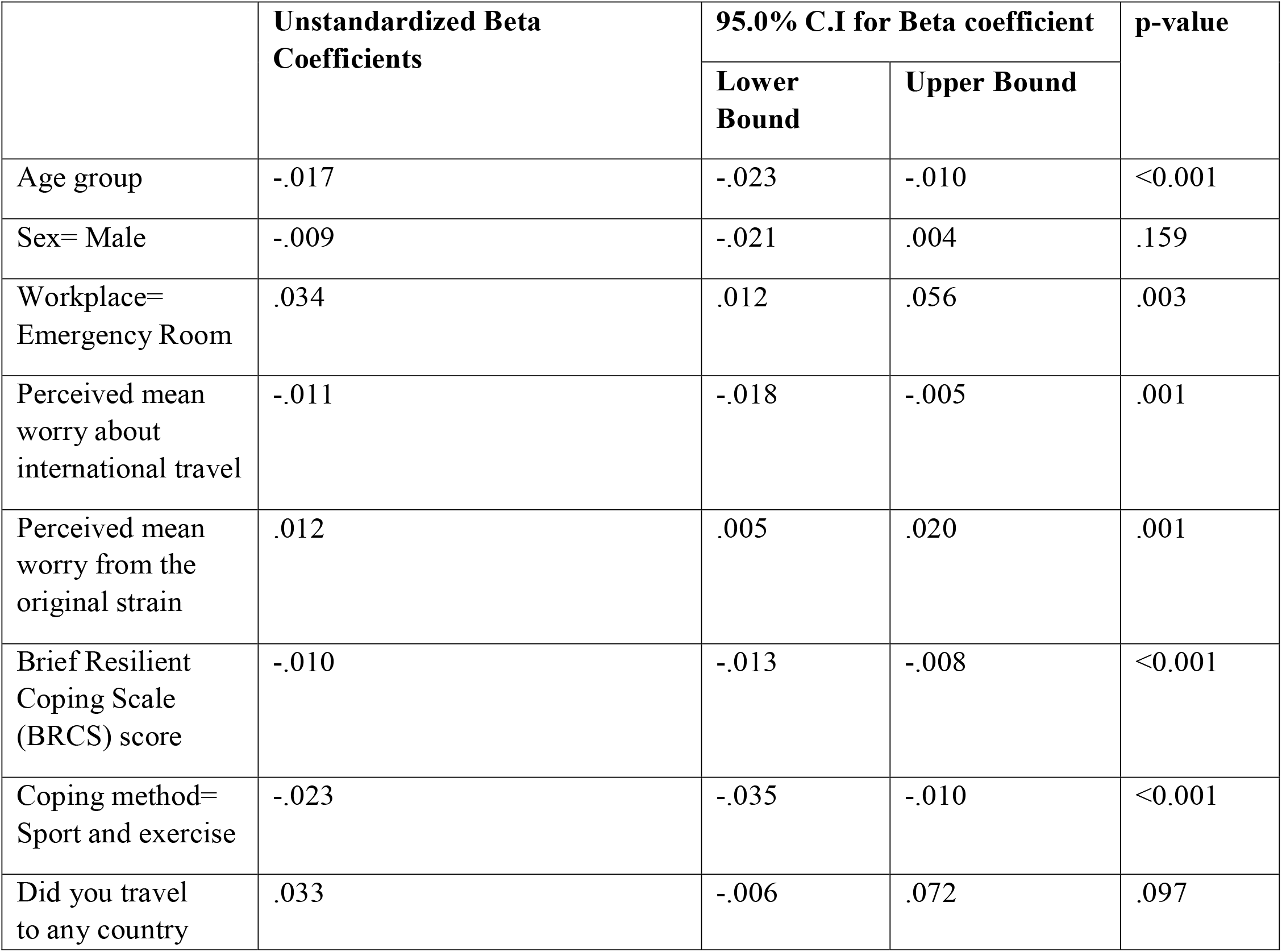
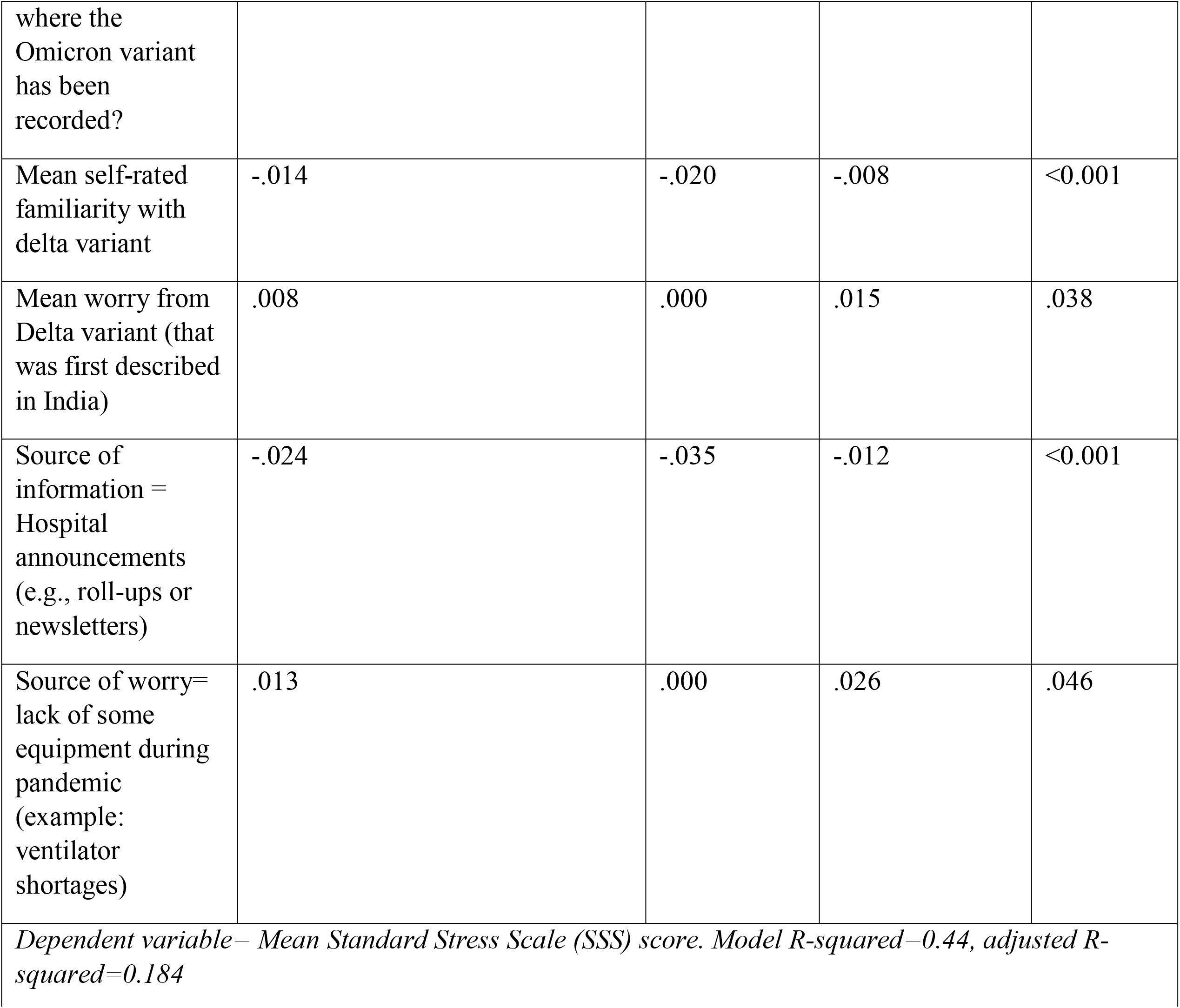
Multivariate Linear Regression Analysis of the HCWs’ stress. N=1285.

Another multiple linear regression model (table 7) was built to explain the variability in HCWs’ stress as scored by the SSS. The model used relevant variables from the survey and is detailed in Table 8. It explained a significant amount of variation in stress scores (adjusted R-squared=0.184, p<0.001). According to this model, increasing resilient coping scores was associated with a significant reduction in stress scores. In other words, an increment of 1 point on the BRCS was associated, on average, with a 0.01 decrement on the SSS, when other variables in the model were held constant. Furthermore, increasing the HCWs’ age groups was associated with a significant decrease in stress scores (see Figure 3). Other variables that were associated with a reduction in stress scores included higher reported worry about international travel, higher self-rated familiarity with the delta variant, coping using sport and exercise, and utilizing hospital announcements as sources of information. Meanwhile, reporting higher levels of worry about the original strain of SARS-CoV-2 or the delta variant was associated with higher stress scores. Similarly, working in the emergency room or reporting that lack of hospital equipment during the pandemic was a source of worry were both associated with higher stress on average.

**Figure 3:**
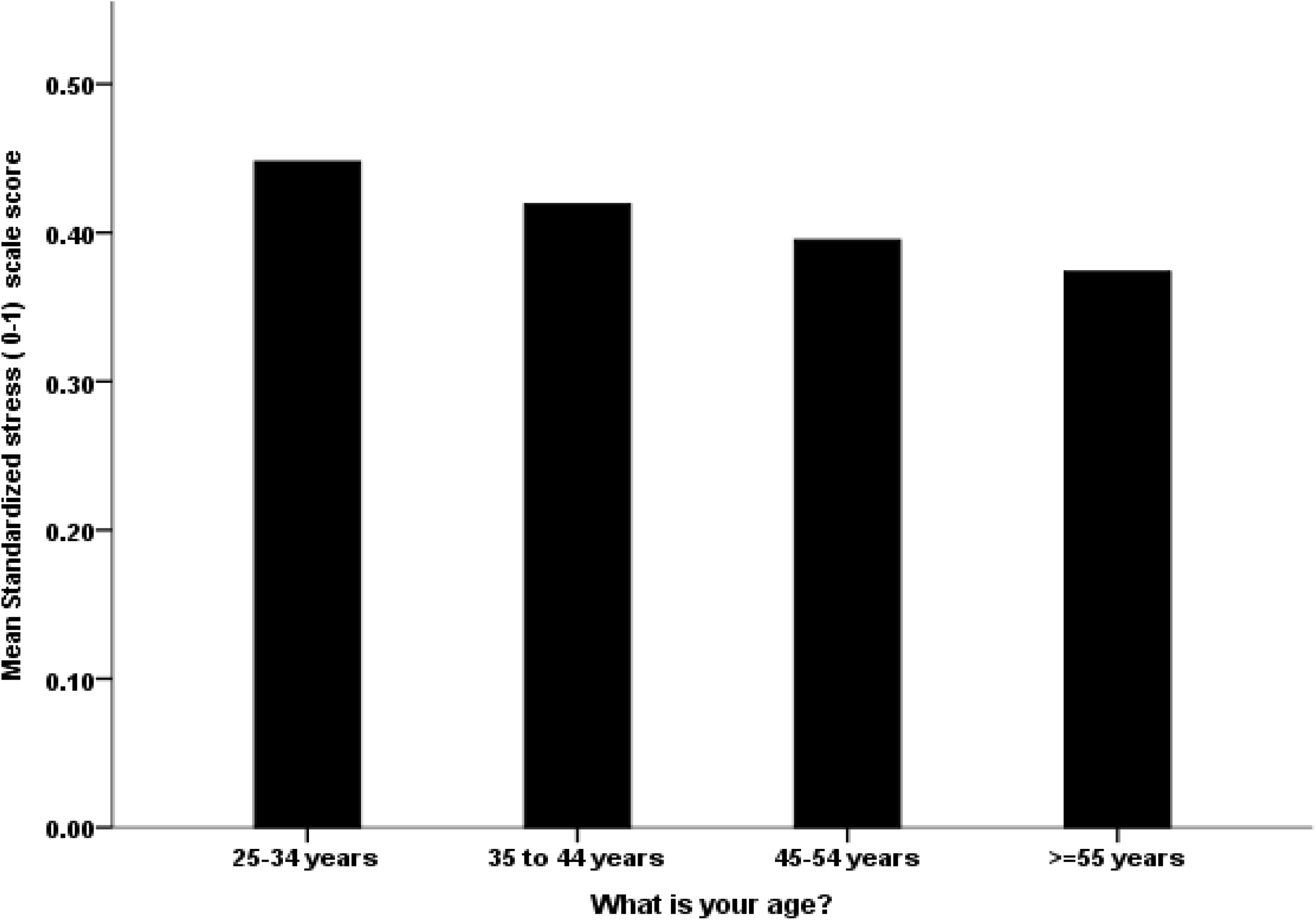
The association between the HCW’s age group with their mean perceived standardized stress score.

## 4 Discussion

To our knowledge, this is the first study to capture the HCWs perceived stress, uncertainty, and coping in the first week of announcing the new Omicron variant. As the picture becomes clearer, two weeks after the announcement of this variant as a variant of concern by WHO, there was a huge public panic evident by economic volatility, and major anticipation and distress of HCWs and healthcare policymakers around the globe [46]. Preliminary data suggested that it might be a more virulent variant, which could lead to a surge of infection and potentially multiple community and household outbreaks. This resulted in calls for declaring extreme caution is required right now and suggesting travel restrictions to be imposed, as well as a race to speed up booster immunization programs and revived efforts to address vaccine inequities [47].

While our study had more female respondents, this is similar to other studies among HCWs that also showed more female gender percentages[37,41]. This could reflect having more female HCWs percentage in the COVID-19 front lines. Women account for 75% of all HCWs worldwide, and they have been disproportionately affected by the pandemic[48]. Furthermore, half of the participants were nurses, with 62.3% being expatriates, making our sample similar to the Saudi Arabian HCW structure, which includes physicians (36.2%) and nurses (63.8%)[49]. Expatriate female nurses in KSA account for most of the nursing workforce, highlighting the need to seek their input and worries more during such a pandemic crisis and international travel restrictions[41,50].

The majority of the HCWs surveyed were frontline HCWs involved in direct patient care, hence the high rate of exposure to COVID-19 positive patients, numbers of COVID-19 PCR tests done and the high rate of prior COVID-19 PCR positivity. And since KSA opened its borders for international travel in Oct 2021, very few HCWs had the chance to travel abroad (2% among the HCWs surveyed). Interestingly, in contrast to a previous surveys conducted among HCWs in KSA about the previous Alpha variant, in which the social media was the highest reported source of information about that variant, in the Omicron setting, the most common source of information about the global and national COVID-19 situation is the WHO and Saudi health authorities, represented by the Saudi CDC and MoH[41].

The number one strategy of HCWs of resilient coping was the belief of growing in positive ways by dealing with difficult situations. In one study from Greece, lack of resilience and the occurrence of maladaptive coping strategies were associated with secondary traumatic stress[51]. Thus, one study created a toolkit for Emotional Coping for Healthcare Staff (TECHS) to provide an online program to help HCWs to cope with the COVID-19 pandemic[52].

The number one reason for intolerance of uncertainty (IUS-12) was that HCWs wanted to be able to organize everything in advance. Generally, HCWs tend to be more on the need for the occurrence of predictability and active engagement to increase certainty and avoid uncertainty[31]. One study showed that maladaptive coping behaviors and being neurotic were associated with risks of distress among HCWs during the COVID-19 epidemics[53].

In this study, the average resilient coping score was 14.31, which is equivalent to 71.6% on the BRCS (as the score ranges from 4-20). And 60.4% of studied HCWs had medium or high resilient coping when categorized by cut-off scores. This indicates that most of the respondents had good resilient coping with the evolving COVID-19 pandemic. Similarly, in a previous study the respondents had a normal range of resilient coping with a mean BRSCLJscore of 14.9[34]. The average Intolerance of Uncertainty Scale (IUS-12) score was 31.8, or 53.0% if the possible score range is changed to a percentage. In a study from Italy, high levels of prospective intolerance of uncertainty as measured by the IUS-12 was a well-recognized feature of the studied Italian HCWs[31]. This is understandable as HCWs tend to have a tendency to be able to predict the future and to relay the prognosis and outcome of disease status to their patients and families.

The mean Standard Stress Scale (SSS) score in our sample of HCWs was 0.42, from a possible range of 0-1. In a previous study from Saudi Arabia, high, moderate and low stress levels were found among 15.8%, 77.2% and 7% of respondents, respectively[6].

The emergence of the Omicron variant had caused increasing worry among HCWs. The top perceived source of concern was the risk of transmitting the infection to household members (63.2%), the risk of the possible implementation of lockdown or disruption of daily life (63.1%) and possible travel ban (61.1%). In a previous study from Saudi Arabia, HCWs were also worried about travel restrictions with the emergence of the B.1.1.7 variant[41]. The current study was done early on after the announcement of the Omicron variant. Since then, many countries announced the ban of travel to and from affected countries such as Southern Africa[54].

Regarding lockdown, a previous study of the B.1.1.7 variant about 53% of surveyed HCWs in Saudi Arabia indicated the worry about lockdown if that variant reached KSA[41]. And another study the response to the worry about lockdown was only thought of if the delta variant would cause a second wave[16]. In the United Kingdom, a third lockdown was done in January 2021 to prevent excess deaths form a third wave[55]. Our findings regarding the relationship between coping and stress emphasize the importance of studying these constructs in times of crisis. This is especially true for HCWs when they are facing stressful situations loaded with uncertainties. Our results show that HCWs who reported better resilient coping during the first week after the announcement of the Omicron variant of COVID-19 had lower stress levels on average. A similar relationship was reported in previous studies on HCWs during the COVID-19 pandemic [34,35]. To the best of our knowledge, this report is the first to demonstrate this relationship during the first days of the emergence of a new variant, highlighting the need for stress management and psychological support for HCWs as the pandemic evolves This work also supports the association between intolerance of uncertainty and negative mental health outcomes, including perceived stress, as has been shown in numerous studies studying both HCWs and the general public during the pandemic[22,23,26,28-30]. Intolerance of uncertainty has been mostly associated with anxiety disorders, especially Generalized Anxiety Disorder (GAD)[56,57], but is likely a trans-diagnostic construct predisposing to many psychiatric disorders [21,58-60]. This suggests that higher intolerance of uncertainty may be an important predictor of psychological distress in HCWs during stressful and uncertain events, such as new infectious threats, as was shown by the relationship between IUS scores and stress in our sample. Our correlation analysis also shows how worries about certain aspects of the new COVID-19 variant were substantially correlated with each other. Studying the sources of anxiety for HCWs and how they relate to each other is important not just for the sake of their wellbeing, but also for the wellbeing of the patients who rely on them during the hardships of the pandemic[61].

Our attempt to explain the differences between the studied HCWs in their reported resilient coping showed that at least some of the differences can be explained by the variables included in the survey such as clinical role. For example, previous studies have reported that nurses may be more likely to report higher stress and anxiety[12,13,62-64] (16–19), in addition to lower coping during the COVID-19 pandemic[34]. These findings may be skewed by the fact that nurses are mostly female, but they could also be explained by the fact that they spend more time on inpatient wards, provide direct patient care, and are in charge of collecting sputum for virus detection, all of which increase their risk of exposure to COVID-19 patients. Furthermore, because of their intimate proximity to patients, they may be more vulnerable to moral injury in the form of suffering, death, and ethical conundrums (20). Similarly, our survey demonstrates how this subgroup of HCWs is more likely to report lower resilient coping when a new variant is discovered. As was reported in a previous study, the relationship between stress and resilient coping explained some of the variability in resilient coping in HCWs [65].

The stress that HCWs reported shortly after the announcement of the Omicron variant varied in our study, and some of that variation could be explained by their underlying intolerance of uncertainty. As has been demonstrated in other HCWs’ samples [31,36], higher intolerance of uncertainty is associated with higher stress. It is likely that this association is particularly relevant when new infectious strains are discovered, since such events are often shrouded with uncertainties. Younger age groups in our sample were significantly more likely to report higher stress. Notably, it has become a consistent finding in COVID-19 research that younger age is associated with worse mental health outcomes in the general public [66]. This finding might be attributed to their function as caregivers in families (particularly females) who provide financial and emotional support to children and the elderly. Remarkably, reporting the use of sport and exercise to cope was associated with a reduction in stress in our sample. This is a finding which may suggest a role for certain healthy coping strategies to help HCWs with the stress of the pandemic, as other authors have suggested [32,65]. Notwithstanding the importance of stress intervention for all HCWs during the pandemic [67,68], our results suggest certain groups are in greater need when new strains of COVID-19 emerge. For example, special attention may be needed for emergency department staff and those worried about the lack of hospital equipment.

### 4.1 Study limitations and strengths

This research is subject to the limitation of cross-sectional studies, including sampling or recall bias possibility and response rate. While our study is among the pioneer research to explore worries and reliance among HCWs considering the novel SARS-CoV-2 Omicron variant, the HCWs stress and perceptions are likely to change as more data about this variant emerges over time. Moreover, HCWs’ coping strategies may differ from one setting to another, so future research could explore this further in other countries.

## 5 Conclusions

This was an early investigation of the correlates of stress and resilient coping of HCWs immediately after the emergence of the Omicron variant of COVID-19 in late 2021. This report demonstrated the inverse relationship between resilient coping and stress, and how underlying intolerance of uncertainty is associated with higher stress in HCWs shortly after the emergence of a new infectious threat. These findings can inspire further research into the mental health of HCWs as the pandemic evolves. Similarly important, our results may help inform policy makers on how to better support front line HCWs in their struggle to perform their duties in uncertain times of new variants outbreaks.

## Data Availability

All data produced in the present study are available upon reasonable request to the corresponding author.

## Abbreviations

BRCS: Brief Resilient Coping Scale
CDC: Centers for Disease Control and Prevention
COVID-19: Coronavirus disease 2019
HCW: healthcare workers
IUS: Intolerance of Uncertainty Scale
MoH: Ministry of Health
SARS-CoV-2: Severe acute respiratory syndrome coronavirus 2
SSS: Standard Stress Scale
WHO: World Health Organization

## Acknowledgments

This research has been financially supported by Prince Abdullah Ben Khalid Celiac Disease Research Chair, under the Vice Deanship of Research Chairs, King Saud University, Riyadh, Kingdom of Saudi Arabia. The research team is thankful for the statistical data analysis consultation offered by www.hodhodata.com.

## Author contributions

MHT, SAlenezi, MA, FA, KA, FA, SAl-Subaie, MB, ZM, and JA conceptualized the study, analyzed the data, and wrote the manuscript.

RA, RB, FA, AAlhaboob, AAlaraj, NSA, RH, LA, NA, AJ, and WA contributed to the study design; collected, analyzed, interpreted data; and edited the manuscript.

All authors reviewed and approved the final version of the manuscript.

---

**Appendix Table.3A:**
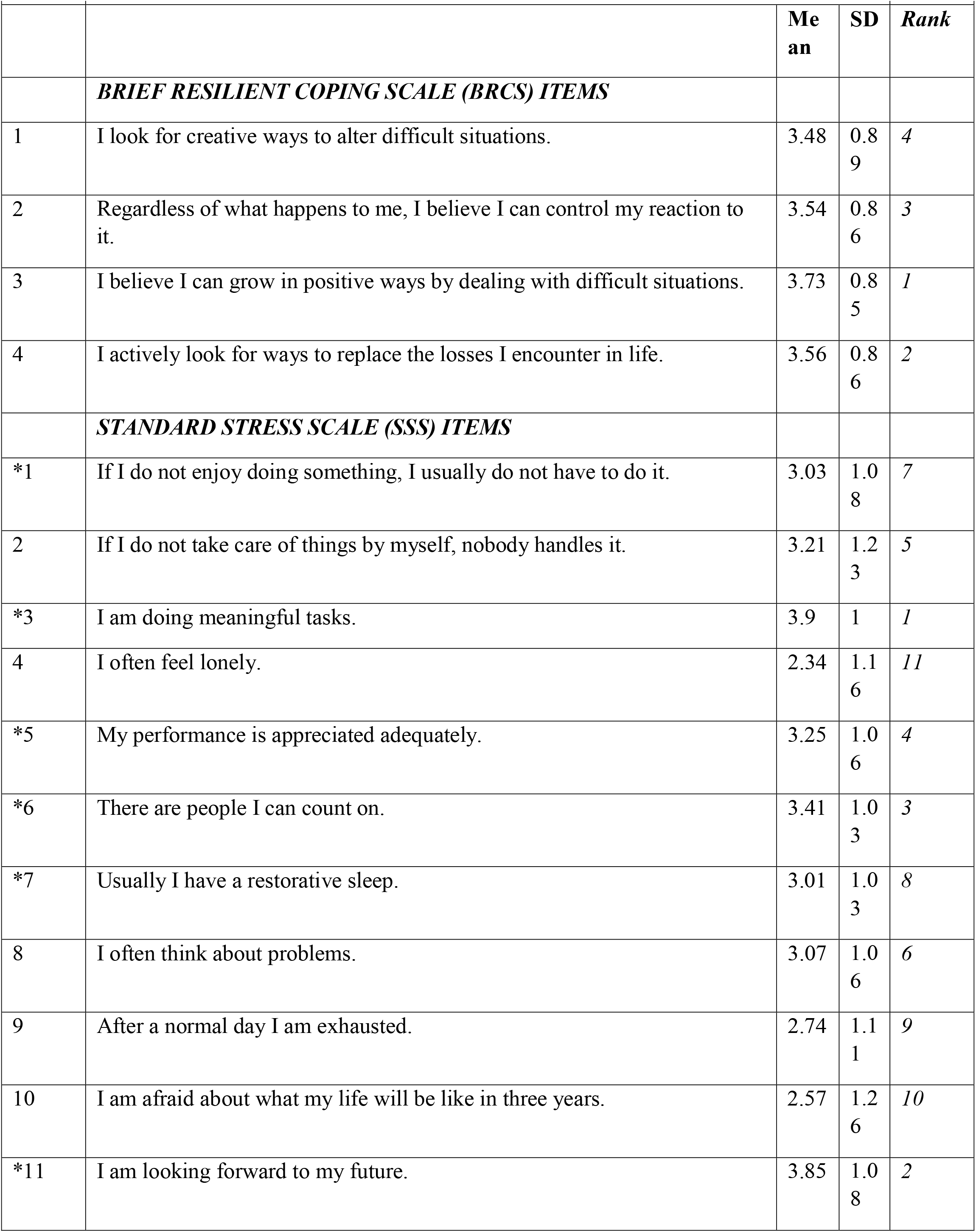
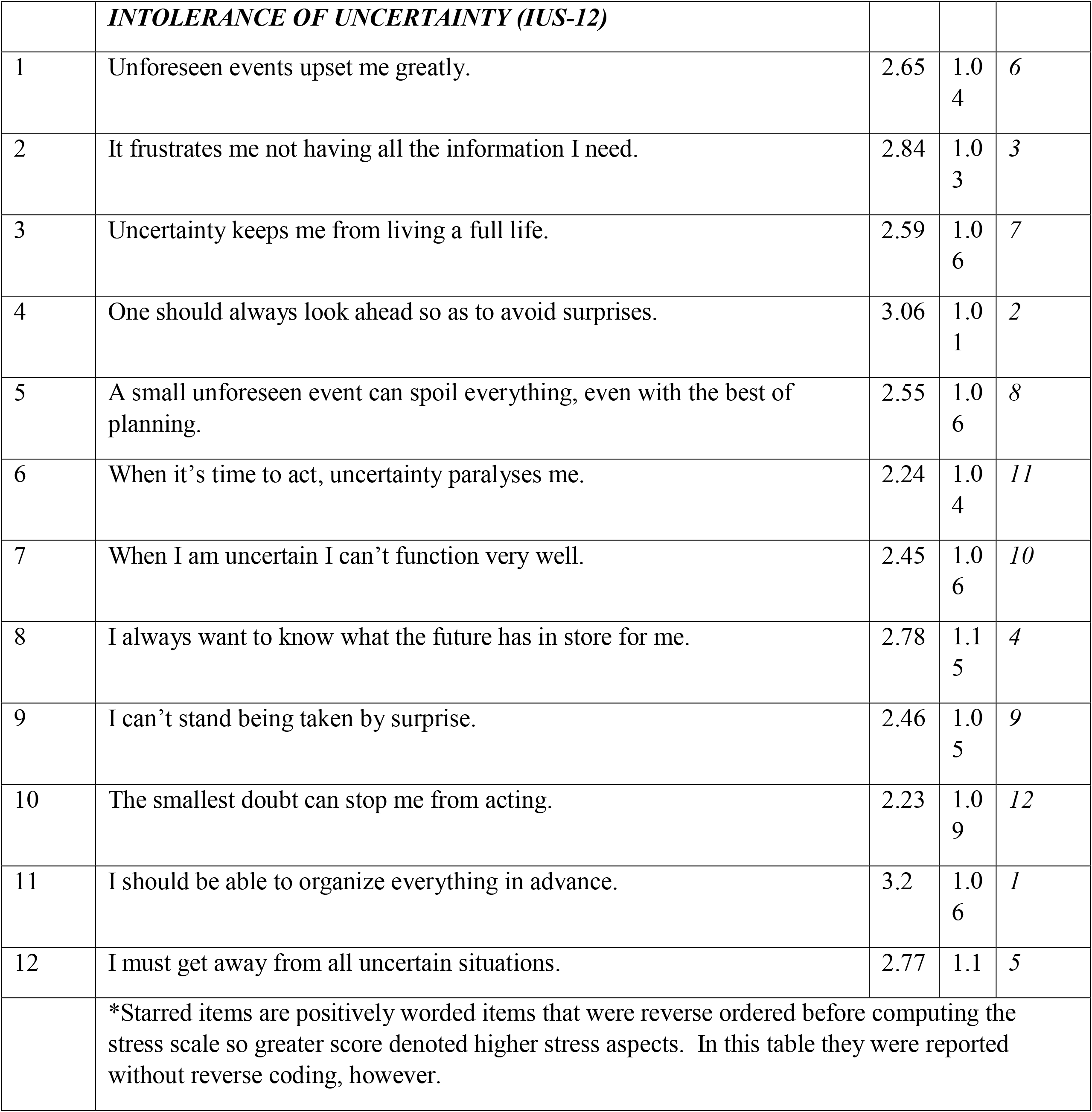
Descriptive analysis of the HCWs’ perceived resilient coping, stress, and intolerance of uncertainty.

|  |  |  |  |  |
| --- | --- | --- | --- | --- |
|  |  | **Me**<br>**an** | **SD** | **Rank** |
| ***BRIEF RESILIENT COPING SCALE (BRCS) ITEMS*** |  |  |  |  |
| 1 | I look for creative ways to alter difficult situations. | 3.48 | 0.89 | 4 |
| 2 | Regardless of what happens to me, I believe I can control my reaction to it. | 3.54 | 0.86 | 3 |
| 3 | I believe I can grow in positive ways by dealing with difficult situations. | 3.73 | 0.85 | 1 |
| 4 | I actively look for ways to replace the losses I encounter in life. | 3.56 | 0.86 | 2 |
| ***STANDARD STRESS SCALE (SSS) ITEMS*** |  |  |  |  |
| *1 | If I do not enjoy doing something, I usually do not have to do it. | 3.03 | 1.08 | 7 |
| 2 | If I do not take care of things by myself, nobody handles it. | 3.21 | 1.23 | 5 |
| *3 | I am doing meaningful tasks. | 3.9 | 1 | 1 |
| 4 | I often feel lonely. | 2.34 | 1.16 | 11 |
| *5 | My performance is appreciated adequately. | 3.25 | 1.06 | 4 |
| *6 | There are people I can count on. | 3.41 | 1.03 | 3 |
| *7 | Usually I have a restorative sleep. | 3.01 | 1.03 | 8 |
| 8 | I often think about problems. | 3.07 | 1.06 | 6 |
| 9 | After a normal day I am exhausted. | 2.74 | 1.11 | 9 |
| 10 | I am afraid about what my life will be like in three years. | 2.57 | 1.26 | 10 |
| *11 | I am looking forward to my future. | 3.85 | 1.08 | 2 |
|  | ***INTOLERANCE OF UNCERTAINTY (IUS-12)*** |  |  |  |
| 1 | Unforeseen events upset me greatly. | 2.65 | 1.0<br>4 | 6 |
| 2 | It frustrates me not having all the information I need. | 2.84 | 1.0<br>3 | 3 |
| 3 | Uncertainty keeps me from living a full life. | 2.59 | 1.0<br>6 | 7 |
| 4 | One should always look ahead so as to avoid surprises. | 3.06 | 1.0<br>1 | 2 |
| 5 | A small unforeseen event can spoil everything, even with the best of planning. | 2.55 | 1.0<br>6 | 8 |
| 6 | When it's time to act, uncertainty paralyzes me. | 2.24 | 1.0<br>4 | 11 |
| 7 | When I am uncertain I can't function very well. | 2.45 | 1.0<br>6 | 10 |
| 8 | I always want to know what the future has in store for me. | 2.78 | 1.1<br>5 | 4 |
| 9 | I can't stand being taken by surprise. | 2.46 | 1.0<br>5 | 9 |
| 10 | The smallest doubt can stop me from acting. | 2.23 | 1.0<br>9 | 12 |
| 11 | I should be able to organize everything in advance. | 3.2 | 1.0<br>6 | 1 |
| 12 | I must get away from all uncertain situations. | 2.77 | 1.1 | 5 |
| *Starred items are positively worded items that were reverse ordered before computing the stress scale so greater score denoted higher stress aspects. In this table they were reported without reverse coding, however. |  |  |  |  |

